# Whole-exome sequencing on 6215 school-aged children reveals the importance of genetic testing in high myopia

**DOI:** 10.1101/2023.06.15.23291239

**Authors:** Xiangyi Yu, Jian Yuan, Kai Li, Yinghao Yao, Shilai Xing, Zhengbo Xue, Yue Zhang, Hui Peng, Gang An, Xiaoguang Yu, the Myopia Associated Genetics and Intervention Consortium, Jia Qu, Jianzhong Su

## Abstract

**Importance:** High myopia (HM) is one of the leading causes of visual impairment and blindness worldwide. It is well-known that genetic factors play a significant role in the development of HM. Early school-aged population-based genetic screening and treatment should be performed to reduce HM complications.

**Objective:** To identify risk variants in a large HM cohort and to examine the implications of universal genetic testing of individuals with HM with respect to clinical decision-making.

**Design, setting, and participants:** In this cross-sectional study, we retrospectively reviewed whole-exome sequencing(WES) results for myopia-related genes in 6,215 school-aged students with HM who underwent germline genetic testing between September 2019 and July 2020. The study setting was a commercial genetic testing laboratory and a multicenter census of elementary and high schools from different educational systems. Participants were aged 6 to 20 years, including 355 primary school students, 1970 junior high school students, and 3890 senior high school students.

**Main outcomes and measures:** The frequency and distribution of positive germline variants and the percentage of individuals with HM (spherical equivalent refraction, SER ≤ -6.00D) in both eyes were detected using the whole-exome sequencing (WES) genetic testing approach.

**Results:** Among individuals with HM, molecular testing yielded 15.52% diagnoses based on systematic analysis of variants in 75 candidate myopic genes. We found 36 known variants in 490 (7.88%) HM cases and 235 protein-truncating variants (PTVs) in 506 (8.14%) HM cases. We found that diagnostic yield was significantly positively associated with SER (*P* = 0.0108), which ranged from 7.66% in the common High Myopia group (HM, -8.00D ≤ SER ≤ -6.00D) to 11.90% in Extreme Myopia group (EM, SER < -10.00D). We also found that primary school students (≤ 11 years) with EM had the highest diagnostic rate of PTV variants (22.86%), which was 1.77 and 4.78 times that of the Ultra Myopia (UM, -10.00D ≤ SER < -8.00D) and HM, respectively.

**Conclusions and relevance:** Using whole-exome sequencing, multiple previously discovered mutations and PTVs which have not been reported to be associated with HM were identified in a substantial number of school-age students with HM. The high mutation frequency in younger students with EM can provide clues for genetic screening and further specific clinical examinations of HM to promote long-term follow-up assessment.

**Key Points:** 

**Question:** Is genetic testing necessary in the diagnosis of hereditary high myopia?

**Findings:** A total of 271 potential pathogenic mutations were identified in 964 of 6,215 (15.52%) students with high myopia (HM) through systematic analysis of variants in 75 candidate genes, including 36 known variants and 235 variants that have not been reported to be associated with HM. Subsequently, whole-exome sequencing on individuals with high myopia grouped by age and degree of refractive error revealed 4.78 times higher protein-truncating variants in primary school children with extreme myopia group (SER < -10.00D, 23.08%) compared with the common high myopia group (-8.00D ≤ SER ≤ -6.00D, 4.78%, *P* = 0.00045). The results suggest that early genetic testing and screening for pathogenic variants is beneficial for young school-aged children with extreme myopia (SER < -10D).

**Meaning:** This study investigated the significance of whole-exome sequencing in the diagnosis of high myopia population. It revealed the genetic cause of high myopia, and is expected to guide future research and clinical diagnosis of high myopia. As part of the treatment and prevention of high myopia, genetic testing can detect the severity of myopia in young children.

## Introduction

Myopia, the most common cause of visual impairment, is a global public health concern due to its increasing prevalence ^1, 2^. High myopia (HM) is generally defined as myopia with a spherical equivalent refractive error of less than or equal to -6.0 diopters (D). HM is a leading cause of impaired vision due to its association with increased risk of serious ocular complications, most notably retinal degeneration ^3, 4^ or even detachment ^5, 6^. The prevalence of HM was reported to be 2-5% in American, Western European, and Australian populations^7^ compared with 4.5-38% in East Asian populations^8–11^. Similar to many other common diseases, HM is a multifactorial disorder, involving interactions between genetic and environmental factors ^12, 13^. Twin and family studies have demonstrated a high heritability for HM, which was estimated to be ∼90% ^14, 15^.

In several early linkage studies and candidate gene studies for myopia, up to 50 loci and genes were identified before the genome-wide association studies (GWAS) era ^16–23^, but these could not be replicated in other related studies. In a GWAS performed using myopia as a dichotomous outcome or refractive error as a quantitative trait, many significant common genetic variants were identified in several loci ^24, 25^, such as 11q24.1 ^26^, 5p15 ^27^, 4q25 ^28^, 13q12 ^29^, 21q22 ^30^, 15q14 ^31^, and 15q25 ^32^. Recently, whole-exome sequencing has revealed variations in a few genes reported to be associated with HM, including *CCDC111* ^33^, *NDUFAF7* ^34^, *P4HA2* ^35^, *SCO2* ^36^, *UNC5D* ^37^, *BSG* ^38^, *ARR3* ^39^, *LOXL3* ^40^, *SLC39A5* ^41^, *LRPAP1* ^42^, *CTSH* ^42^, and *ZNF644* ^43, 44^. On the other hand, HM has also been identified as a symptom of various forms of retinal dystrophies and systemic syndromes caused by several known genes, including *COL2A1* ^45^, *COL11A1* ^46^, *COL9A1* ^47^, and *COL9A2* ^48^ responsible for Stickler syndrome (OMIM #108300, #604841, #614134, #614284) and *FBN1* ^49^ for Marfan syndrome (OMIM #154700). Variants in these genes were detected in a few cases, with screening of additional HM cohorts expected to uncover more variants. WES studies in HM have made significant advancements by identifying new candidate genes for HM and highlighting the critical role of genetic factors in the development of this condition. For instance, a WES study involving 27 families has uncovered 201 candidate mutations that are associated with HM^66^. Therefore, large-scale genetic screening for HM in a larger cohort of school-aged children could provide further insight into the intersecting contributions of biology and the environment.

In this study, we performed WES on 6,215 school-aged children with HM from the Myopia Associated Genetics and Intervention Consortium (MAGIC) cohort ^50^ to identify variants in known HM-related genes and potentially contributory variants in HM genes. This study applied genetic testing to a larger population than previous reports. To our knowledge, this series of multigene germline testing is the largest in a population with HM, highlighting the potential of genetic testing to improve the diagnosis and management and to allow the definition of specific care pathways.

## Methods

### Study design and participants

As part of our ongoing efforts within the MEIS(Myopic Epidemiology and Intervention Study) ^8^ and MAGIC ^50^, an epidemiologic and genetic analysis of refraction-confirmed myopia was conducted among schoolchildren in Wenzhou City, Zhejiang Province, China. For this analysis, we developed a semi-automated vision examination and an information-entry pipeline, online eyesight status information management system (OESIMS) ^51^, which involves manual inspection of automated refractometry data, automatic data import, and collaboration between clinicians and a statistician. From September 2019 to July 2020, a total of 6,215 school-aged Children with HM in both eyes, including 355 primary school students (grades 1-6 in Chinese education system), 1970 junior high school students (grades 7-9), and 3890 senior high school students (grades 10-12), were included in the WES study (eTable 1 in the Supplement). HM is defined as a spherical equivalent refraction (SER) of (sphere + [cylinder/2]) of -6 diopters (D) or less. We further divided the participants into the three groups: High Myopia group (HM, -8.00D ≤ SER ≤ -6.00D), Ultra Myopia group (UM, -10.00D ≤ SER < -8.00D), and Extreme Myopia group (EM, SER < -10.00D). Certified technicians were trained at the Wenzhou Medical University Affiliated Eye Hospital with respect to standard procedures for determining visual acuity (VA) and autorefraction testing. Each school in the district was equipped with an auto refractometer (GoldEye RM-9000) and electronic logarithmic visual chart (GoldEye CM-1900C) by the Wenzhou Municipal Government. These resources were utilized by trained technicians to examine all participants. VA was evaluated using an ‘E-type’ standard logarithmic visual chart at a distance of 5 m. All students underwent non cycloplegic refraction testing using an automated refractometer, followed by complement subjective refraction testing for validation using an ‘E-type’ standard logarithmic visual chart.

The present study was approved by the Ethics Committee of the Wenzhou Medical University Affiliated Eye Hospital (approval numbers Wmu191204 and Wmu191205). Written informed consent conforming to the tenets of the Declaration of Helsinki and following the Guidance of Sample Collection of Human Genetic Diseases (2021SQCJ5721) by the Ministry of Public Health of China was obtained from all participating individuals or their guardians before the study. Participant completion of the self-administered questionnaire was considered informed consent. In order to maintain the confidentiality of the sample/patient IDs used in this research, it is hereby confirmed that the sample/patient IDs have not been disclosed to any individual or entity outside the research group.

### Whole-exome sequencing

A total of 6,215 HM samples were sequenced on Illumina NovaSeq 6000 sequencers at Berry Genomics using the Twist Human Core Exome Kit. All samples were joint called together and were aligned to the consensus human genome sequence build GRCh37/hg19, and BAM files were processed using BWA ^52^ and Sambamba 0.6.6 (https://lomereiter.github.io/sambamba/). Genotype calling was performed using the Genome Analysis Toolkit’s (GATK) HaplotypeCaller and variants were annotated with Ensembl’s Variant Effect Predictor (VEP v.99) ^53, 54^.

### Variant interpretation

A total of 75 genes were included in this study (eTable 2 in the Supplement), including 16 non-syndromic HM genes, 27 genes of eye syndromes associated with HM, and 32 genes of systemic syndromes associated with HM (hereon, we refer to this gene list as HM genes). All HM genes were extracted from OMIM (Online Mendelian Inheritance in Man, https://omim.org/), IMI-Myopia Genetics Report, and other publication evidence.

Variants from 75 genes were selected from the WES dataset of the 6,215 school-aged population with HM. Variants were prefiltered so that only those passing the GATK VQSR (variant quality score recalibration) metric and those located outside of low-complexity regions were remained. Low-certainty variant positions with a genotype depth (DP) < 10 and genotype quality (GQ) < 20 and heterozygous genotype calls with an allele balance >0.8 or <0.2 were ignored. Protein-truncating variants (PTVs) was classified as “frameshift_variant”, “splice_acceptor_variant”, “splice_donor_ variant”, “stop_gained”, or “start_lost” variants. Variant filtering included the following steps. (1) The present analysis focuses on rare variants with presumed large effect sizes. Therefore, we excluded variants that had minor allele frequencies (MAF) higher than 0.005 from three external exome sequence databases (1000 Genomes, NHLBI Exome Sequencing Project (ESP), and gnomAD) and our MAGIC cohort. (2) We excluded variants not consistent with hereditary patterns, which were only one-hit heterozygous variants in autosomal recessive (AR) genes and homozygous variants in autosomal dominant (AD) genes.

### Statistical Analysis

The statistical analyses included all participants for whom data regarding variables of interest were available. The significance of differences between continuous variables was assessed using analysis of variance, and that of differences between categorical variables was assessed using the *χ*^2^ test. A mean comparison of relevant features was conducted using the Student’s t-test or the Wilcoxon signed rank test. All statistical tests were two-sided at *P*<0.05. R software (ver. 3.6.1) was used for analyses (http://www.r-project.org).

## Results

### Cohort information

Of the 6,215 schoolchildren with HM, 2,937 (47.26 %) were female and 3,278 (52.74 %) were male. Their mean age was 14.87 years and 355 participants (5.71%) were primary school students (age ≤ 11 years), 1,970 (31.70%) were junior school students (aged 12 to 14 years), and 3,890 (62.59%) were high school students (age ≥15 years). The mean SER was -7.51D for the right eye and -7.46D for the left eye. Detailed demographic information is presented in eTable 1 in the Supplement.

### Mutational spectrums revealed for known HM related genes

Through the analysis of the exome sequencing data of 6,215 individuals with HM, a total of 271 potential pathogenic variants in 59 of 75 candidate genes were identified in 964 (15.52%) HM cases, including 36 known variants in 490 HM cases and 235 newly identified PTVs in 506 HM cases. Among 36 variants previously reported in 490 cases, 9 variants in 6 HM genes were identified in 34 (0.55%) participants, 10 variants in 8 eye syndrome genes were detected in 125 (2.01%) participants and 17 variants in 9 systemic syndrome genes were found in 337 (5.42%) participants (eTable 3-5 in the Supplement). The most frequently mutated genes were *COL18A1* (c.4318G>A and c.3523_3524del), accounting for 49.18% (241/490) of the diagnoses, respectively (Figure 1A).

**Figure 1.**
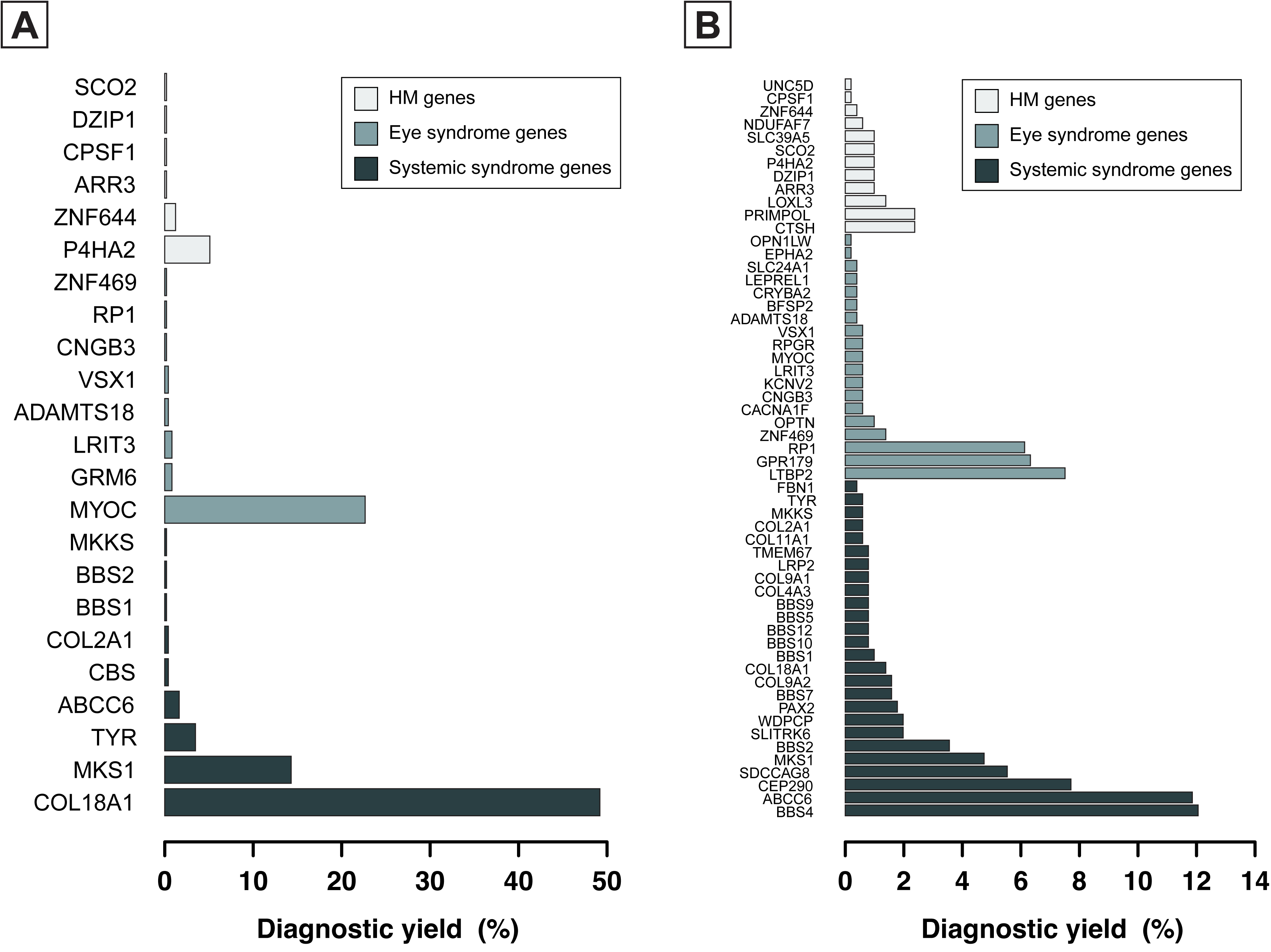
Distribution of mutated genes in the 409 schoolchildren with known variants (A) and 506 schoolchildren with PTVs (B) who received a probable molecular diagnosis after WES genetic testing. Genes without known variants and rare PTVs are not shown.

A total of 235 potentially pathogenic PTVs were identified in 506/6,215 (8.14%) individuals with HM were identified. Among them, 39 PTVs in HM genes were identified in 63/6,215 (1.01%) participants, 75 PTVs of eye syndrome genes in 132/6,215 (2.12%) participants and 121 PTVs of systemic syndrome genes in 327/6,215 (5.26%) participants (eTable 6-8 in the Supplement). Of the 57 genes with PTVs, variants in *BBS4* were the most common, accounting for 12.06% (61/506) of individuals with PTVs (Figure 1B). Of the 235 potentially pathogenic variants, 171 were only found once (i.e., in a single student), whereas 64 occurred in two or more unrelated students, suggesting substantial genetic heterogeneity in HM loci. Variant types included frameshift (85/235), splice acceptor (19/235), splice donor (35/235), stop gained (93/235) and start lost (3/235) variants (Figure 2A). We identified 19 PTVs within *CEP290* and 14 PTVs in *LTBP2* (Figure 2B). Variants in HM-related genes that have not been observed in either public or in-house control populations, including disease-causing model (dominant or recessive) and loss-of-function intolerance (pLI > 0.9). were also detected. They included 14 heterozygous variants in 6 genes (*COL11A1, COL2A1, FBN1, LRP2, UNC5D*, and *ZNF644*) and 2 homozygous mutations in *OPN1LW* and *RPGR* genes (Table 1).

**Figure 2.**
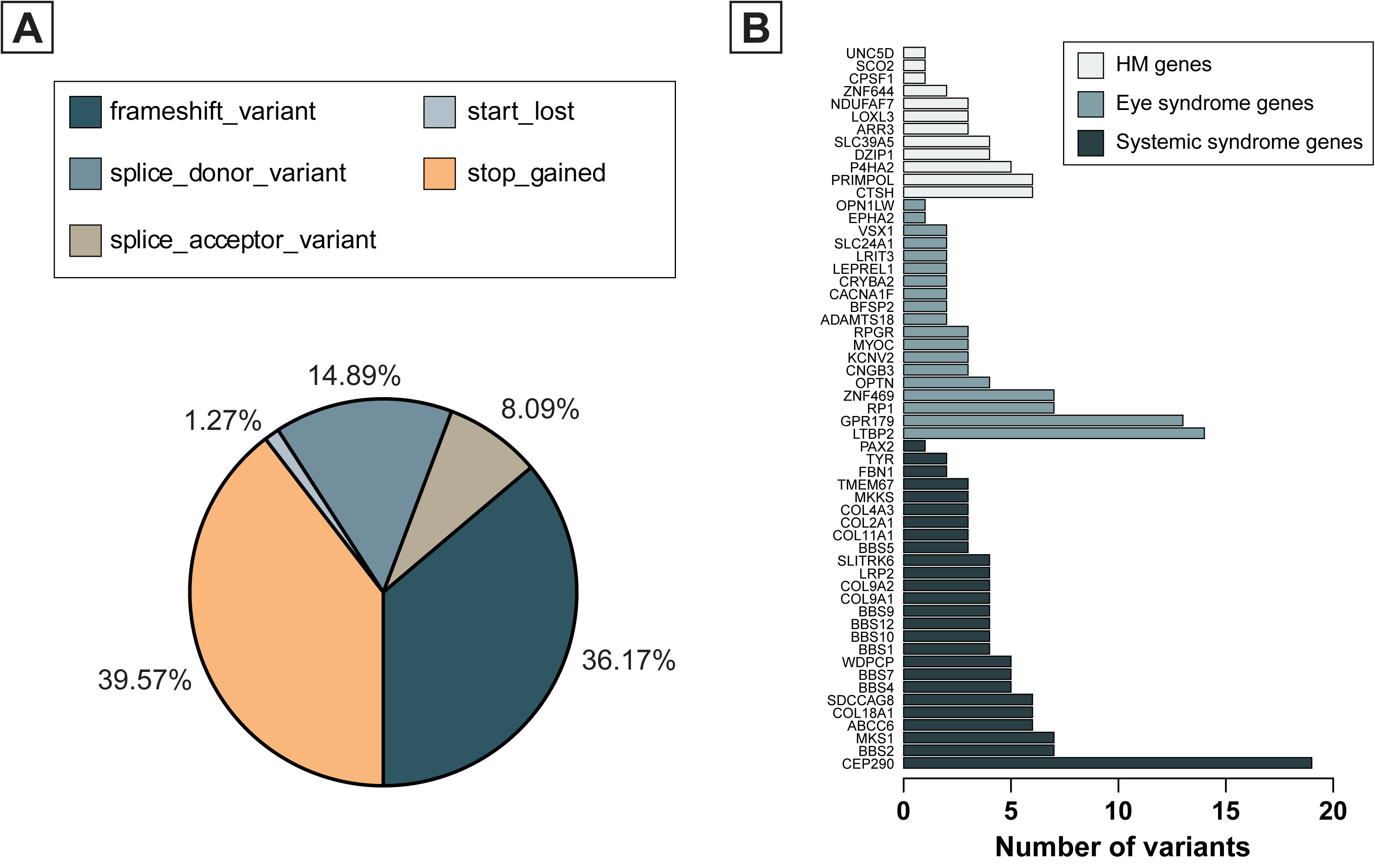
Rare pathogenic variants identified in HM candidate genes. (A) Pathogenic mutation types in HM candidate genes detected in schoolchildren with HM. (B) Number of PTVs identified in HM candidate genes. Barplot is show the frequency of PTV in non-sydromic HM genes, eye syndrome genes and systemic syndrome genes. Genes without rare PTVs are not shown.

**Table 1.** Overview of PTVs in HM candidate genes

| Variant | Gene | Consequences | Transcript | cDNA change | Residue change | LOFTEE | MAF | Status | Sample ID |
| --- | --- | --- | --- | --- | --- | --- | --- | --- | --- |
| chr1_91405539 | ZNF644 | stop_gained | ENST00000370440 | c.1372C>T | p.Arg458Ter | HC | 8.05E-05 | Het | 19BY19988 |
| chr1_91447871 | ZNF644 | frameshift_variant | ENST00000370440 | c.38_39del | p.Lys13IlefsTer2 | HC | 8.05E-05 | Het | 19BY16965 |
| chr1_103449692 | COL11A1 | splice_donor_variant | ENST00000370096 | c.2208+2T>C | - | HC | 8.05E-05 | Het | 19BY08582 |
| chr1_103491076 | COL11A1 | splice_donor_variant | ENST00000370096 | c.897+696G>A | - | HC | 8.05E-05 | Het | 19BY06615 |
| chr2_170002290 | LRP2 | stop_gained | ENST00000263816 | c.12955C>T | p.Arg4319Ter | HC | 8.05E-05 | Het | 19BY04713 |
| chr2_170034316 | LRP2 | frameshift_variant | ENST00000263816 | c.10389dup | p.Ile3464HisfsTer33 | HC | 8.05E-05 | Het | 19BY04553 |
| chr2_170044720 | LRP2 | stop_gained | ENST00000263816 | c.9088C>T | p.Gln3030Ter | HC | 8.05E-05 | Het | 19BY07153 |
| chr2_170063503 | LRP2 | stop_gained | ENST00000263816 | c.6727C>T | p.Arg2243Ter | HC | 8.05E-05 | Het | 19BY11969 |
| chr8_35093311 | UNC5D | frameshift_variant | ENST00000404895 | c.1426G>T | p.Glu476Ter | HC | 8.05E-05 | Het | 19BY02740 |
| chr12_48370913 | COL2A1 | stop_gained | ENST00000380518 | c.3299C>G | p.Ser1100Ter | HC | 8.05E-05 | Het | 19BY04616 |
| chr12_48380199 | COL2A1 | stop_gained | ENST00000380518 | c.1447G>T | p.Gly483Ter | HC | 8.05E-05 | Het | 19BY06672 |
| chr12_48383582 | COL2A1 | stop_gained | ENST00000380518 | c.1030C>T | p.Arg344Ter | HC | 8.05E-05 | Het | 19BY04804 |
| chr15_48829865 | FBN1 | stop_gained | ENST00000316623 | c.679C>T | p.Gln227Ter | HC | 8.05E-05 | Het | 19BY12499 |
| chr15_48829882 | FBN1 | frameshift_variant | ENST00000316623 | c.661del | p.Cys221ValfsTer109 | HC | 8.05E-05 | Het | 19BY06899 |
| chrX_153416181 | OPN1LW | frameshift_variant | ENST00000369951 | c.168del | p.Ser57ValfsTer4 | HC | 1.63E-04 | Hom | 19BY20410 |
| chrX_38176655 | RPGR | frameshift_variant | ENST00000378505 | c.532del | p.Ser178ValfsTer9 | HC | 1.61E-04 | Hom | 19BY06620 |

### Subgroup of cases with extreme myopia preferred to whole-exome sequencing

The analysis revealed that the proportion of cases with PTVs varied across HM conditions and grade groups. The diagnostic yield showed a positive correlation with SER, ranging from 7.66% in the High Myopia group (HM, -8.00D ≤ SER ≤ -6.00D), 8.70% in Ultra Myopia group (UM, -10.00D ≤ SER < - 8.00D) to 11.90% in Extreme Myopia group (EM, SER < -10.00D), with statistically significant increasing trend (Cochran-Armitage test for trend Z = 2.5492, *P* = 0.0108, Figure 3A). The trend was also seen in the independent cohorts of people with HM in either the left or the right eye alone (eFigure 1 in the Supplement). Notably, high SER was significantly associated with a higher diagnostic yield in the primary students (Cochran-Armitage test for trend Z = 3.8848, *P* = 0.0001024) but in other grades (Cochran-Armitage test *P* = 0.1939 for junior school and *P* = 0.3108 for high school) (Figure 3B). The primary students referred for EM had the highest yield (8 of 35 individuals [22.86%]), which was 1.77 and 4.78 times higher than the UM (12.90%, Chi-squared test *P* = 0.1578) HM (4.78%, Chi-squared test *P* = 0.00045), respectively. This trend was consistent across groups divided by the SER of either eye (eFigure 2 in the Supplement). However, for the negative control, the trend was not statistically significant for synonymous variants (eFigure 3 in the Supplement).

**Figure 3.**
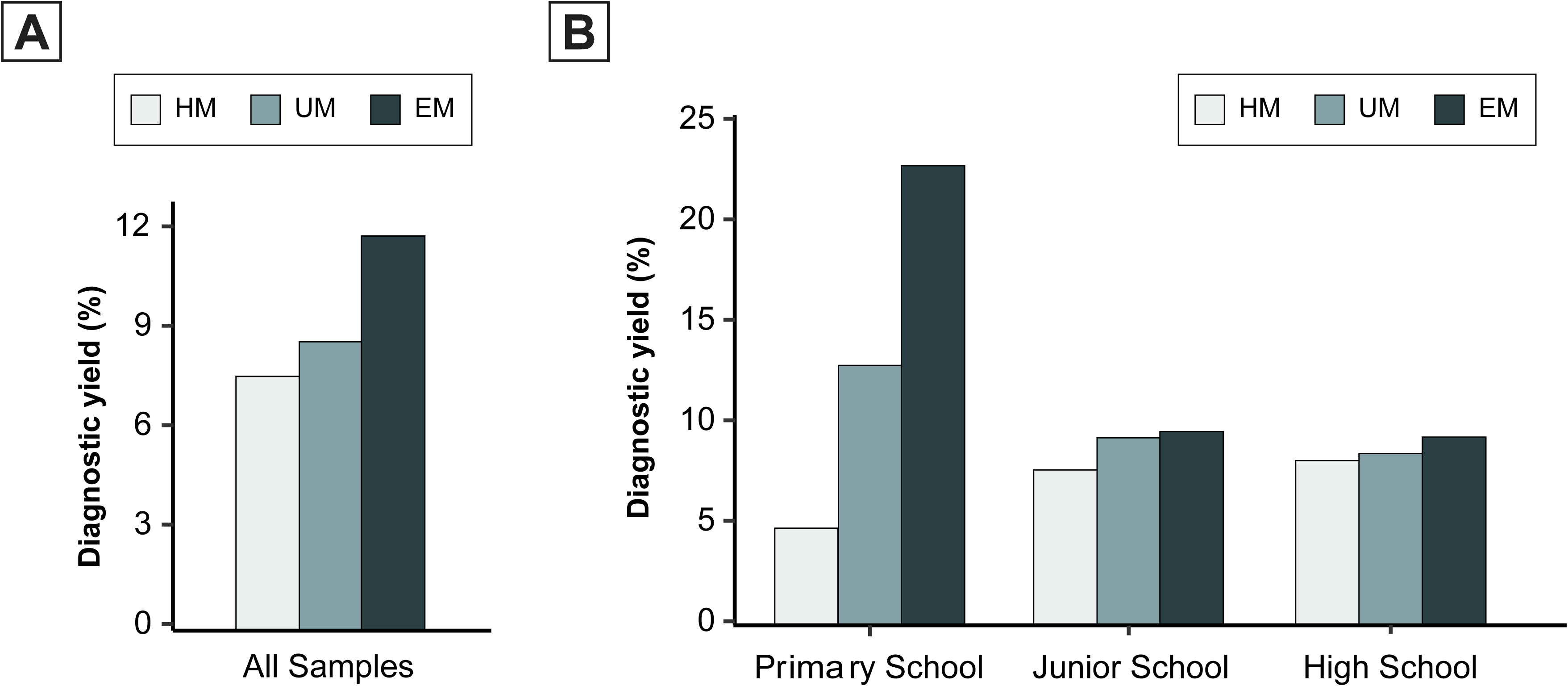
Proportion of schoolchildren with HM carried PTVs in the overall study population (A) and grade relevant subgroups (B). Spherical refraction is range from: HM: -8D ∼ -6D; UM: -10D ∼ -8D; EM: < -10D. The age is calculated by the year who accept the genetic test.

## Discussion

In this study, we tested the diagnostic utility of WES in 6,215 school-aged students with HM. The association of HM with ocular and systemic diseases has been investigated in several previous studies. For example, Marr et al. found that 56% of children with HM had simple symptom of HM, 25% had ocular abnormalities and 19% had systemic disorders ^55^. Another study found mutations in genes known to be involved in retinal disease in about a quarter of the subjects with early-onset HM ^56^. Therefore, 75 genes were included in the present study, including 16 non-syndromic HM genes, 27 genes of eye syndromes, and 32 genes of systemic syndromes associated with HM, such as Stickler syndrome, Marfan syndrome, and Knobloch syndrome.

Systematic analysis of variants in the 75 candidate genes showed that genetic testing had a total diagnostic yield of 15.52%, including 36 known variants in 490 cases and 235 PTVs in 506 cases. The discovery of variants previously obtained in other studies demonstrated the efficiency of genetic approach to find pathogenic variants in our cohort. In our cohort, 36 known variants were found in a total of 490 (7.88%) patients, including a stop-gain variant (NM_004312:c.298C>T; p.Arg100Ter) in *ARR3*^39^ found in one patient. *ARR3* is associated with early-onset HM in a unique X-linked female-limited inheritance ^57^. The most common variant in our study was a missense (NC_000021:c.4318G>A; p.Asp1440Asn) in *COL18A1*, which was detected in 3.86% (240/6215) of our cohort. Mutations in *COL18A1* have been identified in patients with Knobloch syndrome, an inherited disorder characterized by HM, retinal detachment, and occipital defects ^58, 59^.

PTVs result in reduced or abolished protein function and are thus considered to be the most deleterious mutations ^60^.PTVs in HM candidate genes were detected in 506 (8.14%) patients, including *ZNF644*. This is a zinc finger transcription factor expressed in the retina and RPE, which has been suggested to play a role in the development of HM because mutations in *ZNF644* have been found in patients with HM ^43, 61, 62^. In the present study, we found two PTVs in *ZNF644* (c.1372C>T and c.38_39del), which have not been recorded in the gnomAD ^63^. We also found three previously unreported stop-gained variants (c.3299C>G, c.1447G>T, and c.1030C>T) and three splice-donor variants (c.2208+2T>C, c.898-972G>A, and c.897+696G>A) in *COL11A1*. Stickler syndrome is caused by variants of two genes ^64^, and is clinically characterized by HM, which is the most common cause of inherited retinal detachment ^65^. These results indicated that WES could find potential pathogenic variants and provide new genetic evidence for HM candidate genes in our study.

With the increase in the number of high-throughput sequencing projects generating vast amounts of data and the innovation of more statistical methods, the occurrence and development of HM can be systematically explored. The diagnostic efficacy of whole-exome sequencing in the prevention of HM needs to be further evaluated to improve the clinicians and patients’ understanding of this condition and promote its early detection and treatment. A detailed spherical refraction and grade (age)-stratified study showed the pattern of efficiency of genetic testing in different groups. In our study, the detection rate of PTVs increased with the severity of myopia. This trend was further investigated in different age groups. The results showed that only the primary school group had this positive correlation trend between spherical refraction and detection rate. In brief, more severe myopia presentation was found in younger children, suggesting the importance of genetic testing for effective prevention of myopia. Taken together, these findings emphasize the potential value of genetic testing in resolving clinical diagnostic challenges in HM.

## Limitations

Although this study was based on grade (age) and spherical equivalent, detailed ocular and systemic examinations, such as eye axis and fundus photography, were not performed. Therefore, additional ocular biological factors should be considered in future research, which could further provide genetic diagnoses across diverse clinical categories, enabling the identification of novel phenotypic extensions. Furthermore, this study lacks follow-up data over several years, and there is a possibility that HM may progress into ultra or extreme myopia in some patients. A clearer conclusion would be reached if the genetic results could be combined with detailed clinical examinations, especially long-term follow-up assessment for schoolchildren with specific mutations. To ensure data reliability, future multi-year follow-up studies are warranted. In addition, WES currently has limited ability to detect genomic imbalances and does not assess mutations in noncoding regions of the genome, leaving additional blind spots. Physician knowledge of these technical limitations is important as WES is increasingly being incorporated into clinical practice.

## Conclusions

In summary, WES screening can reveal the most important HM candidate genes at once and allows periodic reevaluation of the sequence data to identify new disease genes. In this study, WES showed good diagnostic value for different categories of schoolchildren with HM, yielding the highest diagnosis rate of pathogenic variants in primary school students. Genetic testing is particularly advantageous as it promotes the diagnosis of HM in younger children with more advanced myopia. Our study demonstrated the ability of genetic testing to identify potentially pathogenic variants in HM and reveal new genes contributing to myopia.

## Data Availability

All data produced in the present study are available upon reasonable request to the authors.

## Acknowledgments

We thank Dr. Zhenhui Chen and Dr. Yunlong Ma for their constructive comments regarding this manuscript. We thank Berry Genomics Co., Ltd for sequencing services. This work was supported by the National Natural Science Foundation of China (82172882, U20A20364) and the Zhejiang Provincial Key Research and Development Program Grant (2021C03102) to J. Qu.

## Consortia

The members of the Myopia Associated Genetics and Intervention Consortium (MAGIC) are Jianzhong Su, PhD; Jian Yuan, PhD; Liangde Xu, MD; Shilai Xing, PhD; Mengru Sun, PhD; Yinghao Yao PhD; Fukun Chen, MS; Kai Li, MS; Xiangyi Yu, MS; Zhengbo Xue, PhD; Yaru Zhang, PhD; Ji Zhang, MS; Hui Liu, PhD; Dandan Fan, PhD; Guosi Zhang, MS; Hong Wang, MD; Meng Zhou, PhD; Hao Chen, MD; Fan Lyu, MD; Gang An, MD; Yuanchao Xue, PhD; Zhenhui Chen, PhD; Jian Yang, PhD; Jia Qu, MD; Yunlong Ma, PhD; Yichun Xiong, PhD; Xinting Liu, MD; Nan Wu, MD; Jie Sun, PhD; Jinhua Bao, MD; Liang Xu, MD; Ling Li, PhD; Liang Ye, MD; Jun Jiang, MD; Xinjie Mao, MD; Xinping Yu, MD; Xiaoming Huang, MD; Jingjing Xu, MD; Miaomiao Li, MD; Xuemei Zhang, MD; Liang Hu, MD; Zhuopao Zuo, MD; Wanqing Jin, PhD; Jiawei Zhou, PhD; Yuwen Wang, MD; Xue Li, MD; Fang Hou, MD; Fei Qiu, MS; Yijun Zhou, MS; Na Gao, PhD; Xinyu Wang, PhD; Xinrui Shi, PhD; Yuchun Deng, PhD; Xiaoguang Yu, PhD; Yu Bai, MS; Chenghao Li, MS; Lu Chen, MS; Ke Li, MS; Lijun Dai, MS; Peng Lin, MS; Jingting Zhao, MS; Congcong Yan, MS; Siqi Bao, MS; Zicheng Zhang, MS; Shen Wang, MS; Haojun Sun, MS; Siyi Jiang, MS; Wei Dai, MS; Hengte Kong, MS; Xiaoyan Lu, MS; Jing Li, PhD; Liansheng Li, MS; Siyu Wang, MS.

## Author contributions

The study was conceived, designed, and supervised by J.Q., J.S. and J.Y. Analysis of data was performed by X.Y., J.Y., K.L., S.X., Y.Y., Y.Z., Z.X., G.A. and X.Y. The manuscript was written by X.Y., J.S. and J.Y.

## Data availability

Individual-level data are not publicly available due to ethical and legal restrictions related to the Wenzhou Medical University. Supporting data are available from the corresponding author upon reasonable request but access to data must be granted by the MAGIC committees. VCF files will be uploaded to Genome Variation Map (GVM) in BIG Data Center (http://bigd.big.ac.cn/gsa), Beijing Institute of Genomics (BIG), Chinese Academy of Sciences, with an accession number GVM000296. The datasets used and analyzed during the current study are available from the corresponding author on reasonable request.

