## supplemental figure for "Whole-exome sequencing on 6215 school-aged children reveals the importance of genetic testing in high myopia": SM_Figures_2023May26.docx

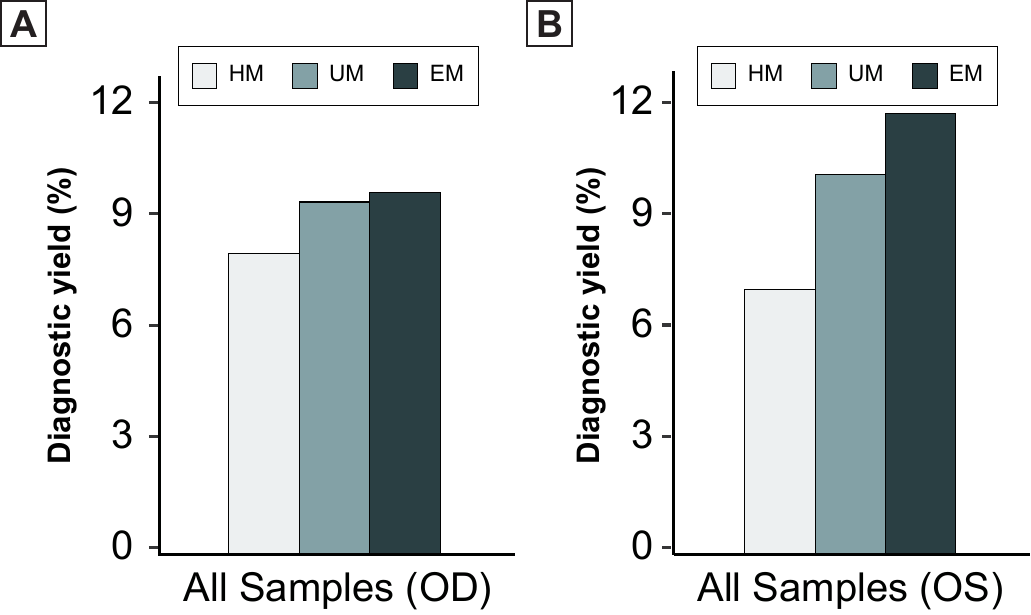

eFigure 1. Proportion of HM cases carried rare PTVs in overall study population. (A) OD: right eyes. (B) OS: left eyes.

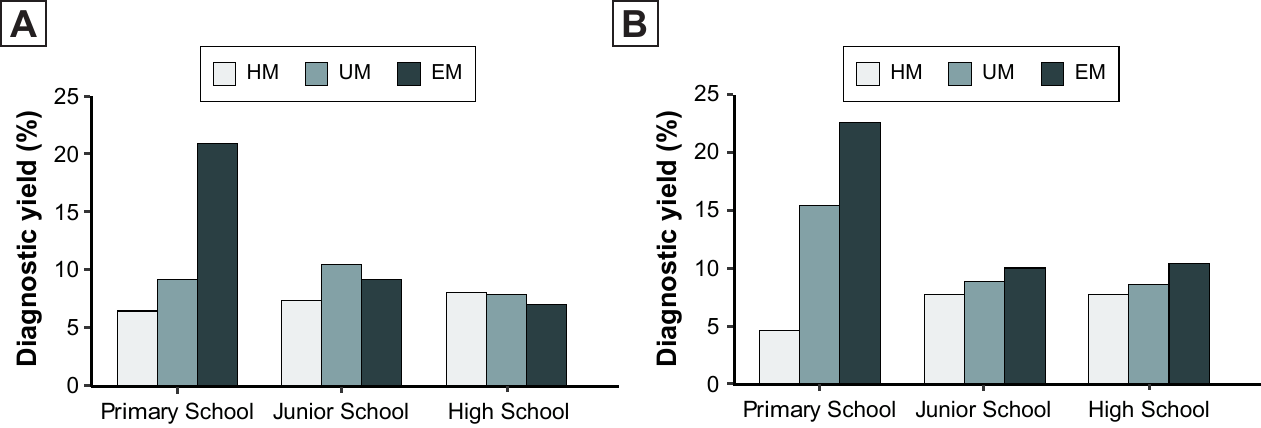

eFigure 2. Proportion of HM cases carried rare PTVs in primary school, junior school, high school. The age is calculated by the year who accept the genetic test. (A) OD: right eyes. (B) OS: left eyes.

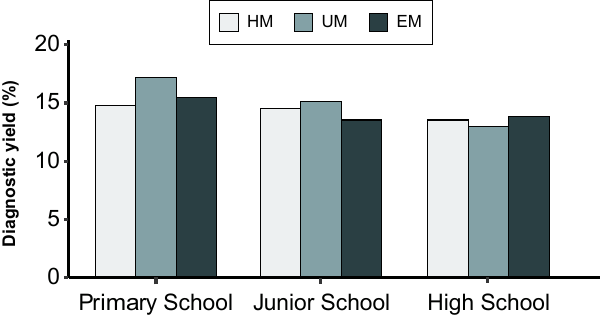

eFigure 3. Proportion of HM cases carried rare synonymous variants in primary school, junior school, high school. The age is calculated by the year who accept the genetic test.
